# *TNF* promoter hypomethylation is associated with mucosal inflammation in IBD and anti-TNF response

**DOI:** 10.1101/2024.02.05.24302343

**Authors:** Daniel S. Levic, Donna Niedzwiecki, Apoorva Kandakatla, Norah S. Karlovich, Arjun Juneja, Jieun Park, Christina Stolarchuk, Shanté Adams, Jason R. Willer, Matthew R. Schaner, Grace Lian, Caroline Beasley, Lindsay Marjoram, Ann D. Flynn, John F. Valentine, Jane E. Onken, Shehzad Z. Sheikh, Erica E. Davis, Kimberley J. Evason, Katherine S. Garman, Michel Bagnat

## Abstract

**Background and aims:** Inflammatory Bowel Diseases (IBD) are chronic inflammatory conditions influenced heavily by environmental factors. DNA methylation is a form of epigenetic regulation linking environmental stimuli to gene expression changes and inflammation. Here, we investigated how DNA methylation of the *TNF* promoter differs between inflamed and uninflamed mucosa of IBD patients, including anti-TNF responders and non-responders.

**Methods:** We obtained mucosal biopsies from 200 participants (133 IBD and 67 controls) and analyzed *TNF* promoter methylation using bisulfite sequencing, comparing inflamed with uninflamed segments, in addition to paired inflamed/uninflamed samples from individual patients. We conducted similar analyses on purified intestinal epithelial cells from bowel resections. We also compared *TNF* methylation levels of inflamed and uninflamed mucosa from a separate cohort of 15 anti-TNF responders and 17 non-responders. Finally, we sequenced DNA methyltransferase genes to identify rare variants in IBD patients and functionally tested them using rescue experiments in a zebrafish genetic model of DNA methylation deficiency.

**Results:** *TNF* promoter methylation levels were decreased in inflamed mucosa of IBD patients and correlated with disease severity. Isolated IECs from inflamed tissue showed proportional decreases in *TNF* methylation. Anti-TNF non-responders showed lower levels of *TNF* methylation than responders in uninflamed mucosa. Our sequencing analysis revealed two missense variants in *DNMT1*, one of which had reduced function *in vivo*.

**Conclusions:** Our study reveals an association of *TNF* promoter hypomethylation with mucosal inflammation, suggesting that IBD patients may be particularly sensitive to inflammatory environmental insults affecting DNA methylation. Together, our analyses indicate that *TNF* promoter methylation analysis may aid in the characterization of IBD status and evaluation of anti-TNF therapy response.

## Introduction

Crohn’s disease (CD) and ulcerative colitis (UC) are complex and heterogeneous chronic inflammatory bowel diseases (IBD) characterized by cycles of severe, relapsing intestinal inflammation. IBDs are complex disorders of poorly understood origin and are generally thought to result from environmental triggers, such as diet and the microbiome, in genetically predisposed individuals. Large-scale association studies have revealed remarkable genetic complexity and identified more than 240 risk loci for IBD^1–3^, although estimates indicate that genetic contributions account for less than 15% of IBD cases^1, 2^. Nevertheless, while there may be many distinct causes of IBD, clinical manifestation (i.e. inflammation) is largely mediated by increased expression of a few key cytokines^4^. Among these, tumor necrosis factor (TNF) is strongly linked to pathogenesis of IBD^5^. Monoclonal antibodies targeting TNF (anti-TNFs) were the first biologic therapeutic agent approved to treat IBD and remain the most effective therapy for achieving endoscopic healing in IBD patients^6^. Additionally, mucosal TNF expression levels closely correlate with disease activity in IBD patients^7^. Therefore, it stands to reason that identifying factors that control TNF levels are critical for understanding the pathophysiology of IBD.

Epigenetics has emerged as a link between environmental triggers, gene expression changes, and aberrant inflammation^8^. Epigenetic modifications, such as DNA and histone methylation, are strongly influenced by environmental stimuli and can regulate gene expression levels^9^. Epigenome wide association studies have identified many differentially methylated loci associated with IBD^10–19^. Additionally, changes in DNA methylation of specific genes have been proposed to mediate some of the known genetic risks for IBD^15^. Stable epigenetic changes specific to CD or UC patients also show promise as potential biomarkers to differentiate IBD subtypes^18, 20^.

We previously found that loss of the DNA methyltransferase gene, *dnmt1*, triggered spontaneous intestinal inflammation and an IBD-like phenotype in zebrafish. This phenotype was mediated by impaired *tnf* promoter methylation and de-repression of *tnf* in intestinal epithelial cells (IECs). Moreover, hypomethylation and overexpression of *tnf* in IECs occurred prior to other inflammatory phenotypes, such as immune cell infiltration^21^. Similar intestinal phenotypes were later observed in mice lacking *Dnmt1* and *Dnmt3b* in IECs^22^. Furthermore, both polymorphisms and decreased expression levels of *DNMT3A* are risk factors for IBD^23^. Together, these studies indicate a critical role for DNA methylation in IECs to suppress inflammation and protect against IBD.

Given the central role of *TNF* in IBD pathophysiology and its importance as a therapeutic target, we investigated whether DNA methylation of its promoter sequence is altered in intestinal mucosa of CD and UC patients using a case-control study of individuals undergoing colonoscopy for IBD disease activity assessment or controls undergoing screening or diagnostic colonoscopy without colitis (n=200 total). *TNF* methylation levels were decreased in both CD and UC patients compared to healthy controls. Importantly, these changes were present only within inflamed mucosa, and *TNF* methylation levels correlated with the severity of local inflammation. Using an independent cohort, we found that *TNF* promoter methylation levels differed according to anti-TNF therapy response, with non-responders also exhibiting hypomethylation in uninflamed mucosa. Finally, we also evaluated whether variants in DNA methyltransferase genes could account for any of the observed methylation changes in our cohort. While our data shows that causal variants in these genes are likely very rare among IBD patients, we identified a missense variant affecting the catalytic domain of DNMT1 that showed reduced activity using *in vivo* assays in zebrafish. Overall, our study indicates that intestinal mucosal inflammation is associated with *TNF* promoter hypomethylation in IBD patients, especially in anti-TNF non-responders. *TNF* methylation may represent a safeguard to dampen the inflammatory response in the intestine.

## Results

### Cohorts

A total of 200 participants were included in the initial phase of this study. Baseline characteristics of the cohort are shown in Table 1. Among the patients with IBD, 86 cases had a diagnosis of CD (64.7%), 46 cases had a diagnosis of UC (34.6%), and 1 case was of indeterminate type. Duration of disease was estimated for all cases with a median of 16 years since initial diagnosis and range of 0-46 years of disease duration. A family history of IBD was present in 38 patients (29.7%).

**Table 1.** Demographics of inflammatory bowel disease cases and screening colonoscopy controls.

|  | **IBD Cases**<br>**(n=133)** | **Controls**<br>**(n=67)** | **All**<br>**(n=200)** | **p-value** |
| --- | --- | --- | --- | --- |
| Age (years) |  |  |  |  |
| Median | 48.3 | 56.1 | 52.0 | <0.001 |
| Range | 21.2-over 80 | 31.8-75.5 | 21.2-over 80 |  |
| Sex |  |  |  |  |
| Female | 78 (58.6%) | 36 (53.7%) | 114 (57.04%) | 0.51 |
| Race/Ethnicity |  |  |  |  |
| Black | 13 (9.8%) | 13 (19.4%) | 26 (13.1%) | 0.12 |
| White | 112 (84.8%) | 49 (73.1%) | 161 (80.9%) |  |
| Other | 7 (5.3%) | 5 (7.5%) | 12 (6.04%) |  |
| Unknown | 1 | 0 | 1 |  |
| Hispanic* | 1 | 1 | 2 |  |

\*Counted in Other Race/Ethnicity category

Among cases, 72 (54.1%) had endoscopic evidence of disease activity at the time of colonoscopy and 91 (68.4%) had histologic evidence of disease. Regarding exposures, 49 cases (37.1%) had any tobacco use, 7 (5.3%) were actively using tobacco at the time of index procedure, 94 cases (71.2%) had any alcohol use, and 72 (54.5%) had current alcohol use.

A prior history of anti-TNF-alpha therapy had occurred in 80 cases (60.1%). Of these, 60 (74.14%) received infliximab as initial therapy. At the time of index procedure, 21 (51.2%) patients were being treated with infliximab and 18 were being treated with adalimumab (43.9%) while one patient was receiving certolizumab (2.4%). Alternatively, 5 patients (3.8%) were being treated with ustekinumab, 11 with vedolizumab (8.4%), and 14 (10.7%) with steroids at the time of index procedure.

### *TNF* hypomethylation is associated with inflammation in CD and UC

To investigate whether *TNF* methylation is altered in IBD patients, we focused on a cluster of CpG sites in the distal promoter (Fig. 1A) that was previously shown to be highly and stably methylated in macrophages from healthy human donors^24^. We first obtained colonic mucosal biopsies from 6 healthy donors and found that 3 distal CpG sites (–304, –245, and –239 bp upstream from TNF exon 1) were highly methylated, whereas the more proximal 8 CpG sites had much lower methylation levels, similar to previous reports^24^ (Fig. 1B, C).

**Figure 1.**
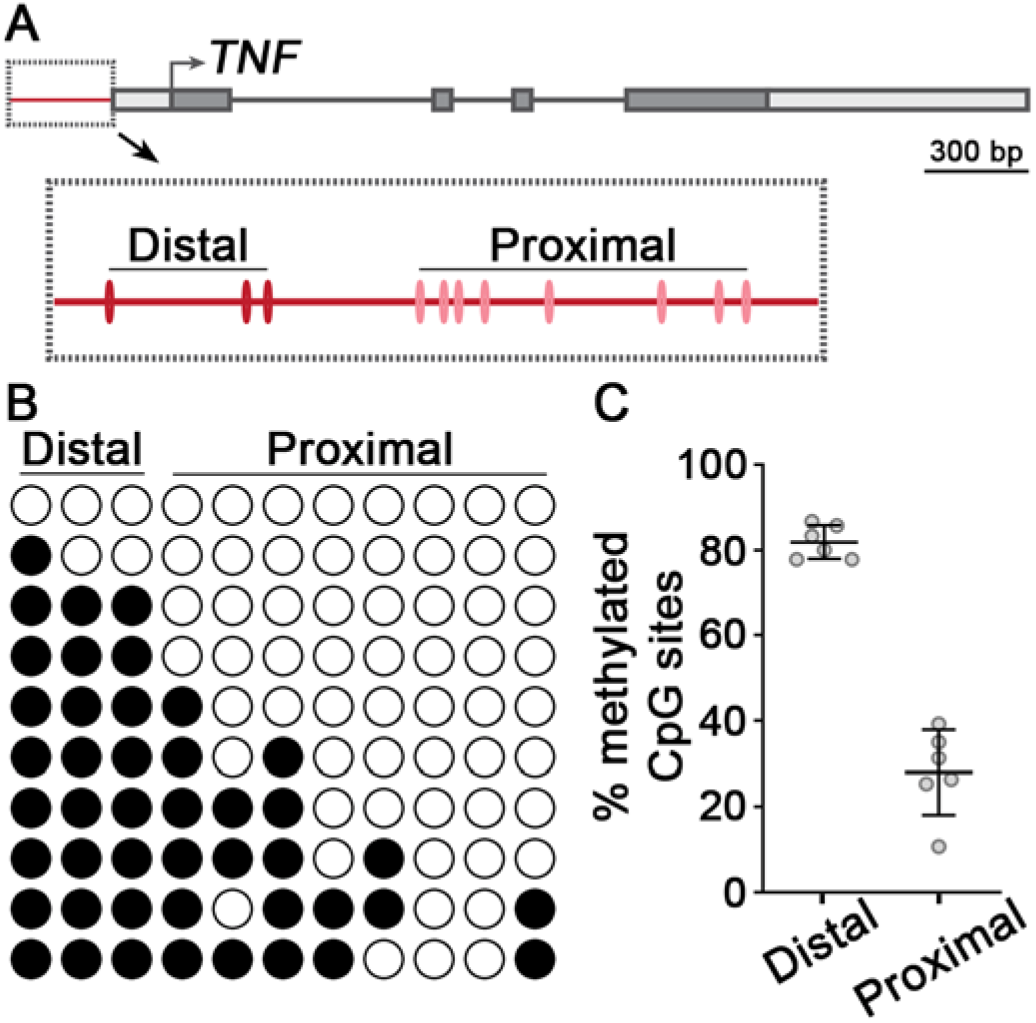
Methylation analysis of the *TNF* promoter from healthy human colon mucosa. (**A**) Schematic of the human *TNF* gene. CpG sites of the promoter region (red line) are shown in the boxed inset. **(B)** Methylation analysis of the *TNF* promoter from healthy human colon using targeted bisulfite sequencing. Columns represent individual CpG sites, and rows are 10 sequencing replicates for a single donor. Filled circles are methylated CpG sites, while open circles are non-methylated. **(C)** Quantification of methylation analysis from healthy human colon. Individual data points are average methylation values from n=6 individual donors. Mean ± SD are plotted. P value was calculated using a non-parametric Wilcoxon two sample test.

We next compared mucosal *TNF* methylation levels of IBD cases (n=199 biopsies from 133 patients) and healthy controls (n=69 biopsies from 67 donors). Among IBD cases, 87 samples were inflamed and 112 were uninflamed; 58 of the uninflamed samples were paired biopsies collected along with inflamed samples. Without accounting for disease activity, no differences were observed between cases and controls (non-parametric, Wilcoxon Two Sample test; p=0.4063) (Fig. 2A). By contrast, when cases were compared according to inflammation status, significant differences were found. *TNF* methylation was higher in uninflamed samples from IBD cases versus inflamed samples (non-parametric, Wilcoxon Two Sample test; p<0.0001) (Fig. 2B), and methylation levels of uninflamed samples from both CD and UC cases were more similar to healthy controls than inflamed samples (Fig. 2C). We validated these findings using an independent cohort of uninflamed and inflamed mucosal samples from CD cases (n=26 biopsies from 15 patients) obtained from a separate study site (non-parametric, Wilcoxon Two Sample test, p=0.0406) (Fig. 2D).

**Figure 2.**
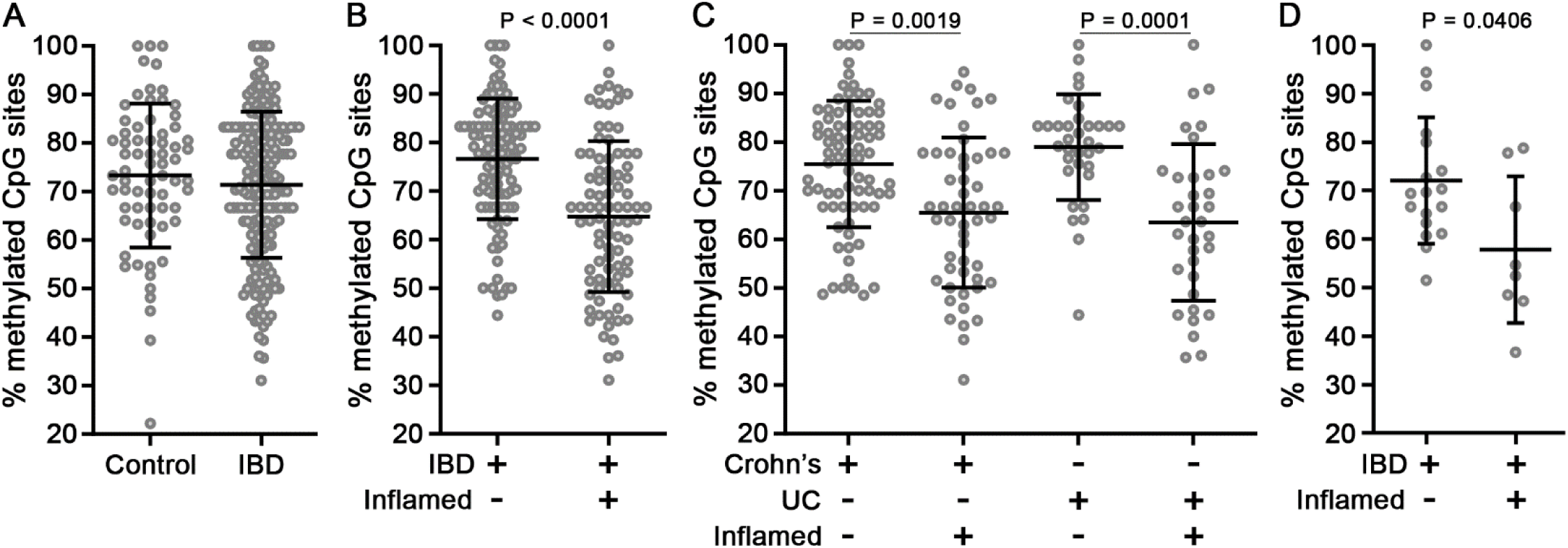
Inflamed mucosa of IBD cases exhibit *TNF* hypomethylation. (**A**) Methylation analysis of the *TNF* promoter from mucosa of controls (n=67 donors; 69 samples) and IBD cases (n=133 donors; 199 samples). **(B)** IBD cases were stratified according to biopsy inflammation status. **(C)** IBD cases were stratified according to disease subtype and biopsy inflammation status. **(D)** Methylation analysis of an independent cohort of IBD cases (n=15 donors; 26 samples) obtained from a separate study site. Data points are average methylation values for individual biopsies. Mean ± SD are plotted. P values were calculated using non-parametric Wilcoxon two sample tests (A, B, D) or a non-parametric Kruskal-Wallis H test (C).

Because inflamed biopsies from cases exhibited *TNF* hypomethylation, we hypothesized that the severity of disease activity might influence methylation levels. To explore this possibility, we analyzed *TNF* methylation levels in uninflamed samples from cases with or without active disease compared to inflamed samples of varying severity. Median *TNF* methylation levels were negatively associated with the degree of disease activity (p <0.0001, Kruskal-Wallis test) (Fig. 3A). To further investigate the relationship between inflammation and *TNF* methylation, we analyzed paired inflamed and uninflamed biopsies from individual cases with active disease (n=58 cases). Inflamed samples were significantly hypomethylated relative to their paired uninflamed samples (non-parametric, Wilcoxon matched-pairs signed rank test, p=0.0001) (Fig. 3B). Although inflamed samples of both groups showed decreases in *TNF* methylation, the differences were significantly more pronounced in UC than CD cases (non-parametric, Wilcoxon matched-pairs signed rank test; UC, Δ=-22.03%, p=0.0002; CD, Δ=-7.89%, p=0.0832).

**Figure 3.**
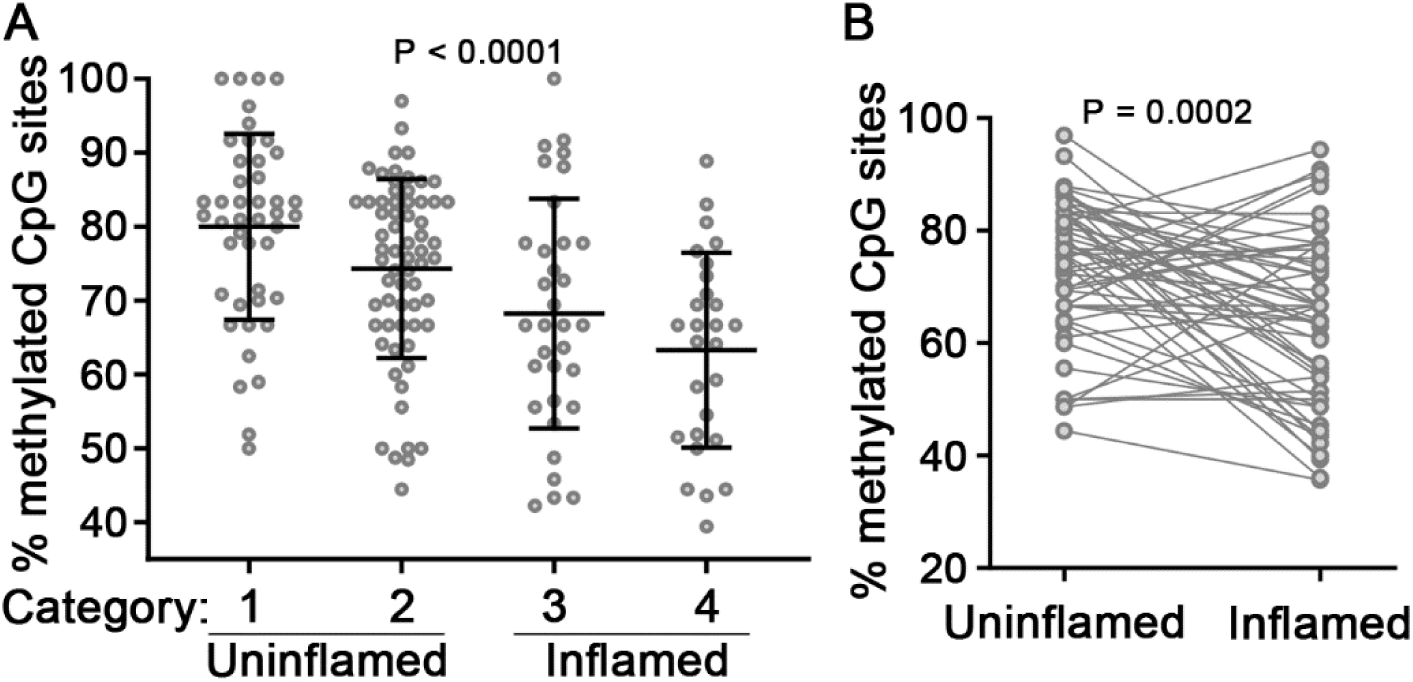
Association of *TNF* hypomethylation with mucosal inflammation. (**A**) *TNF* Methylation analysis of IBD cases stratified by local mucosal disease activity. Categories: 1, uninflamed biopsies from IBD p tients without active disease; 2, uninflamed biopsies from IBD patients with active disease; 3, inflamed biopsies from IBD patients exhibiting mild or focal inflammation; 4, inflamed biopsies from IBD patients exhibiting chronic or acute colitis. Data points are average methylation values for individual biopsies. Mean ± SD are plotted. **(B)** *TNF* Methylation analysis of paired uninflamed and inflamed mucosal samples from individual IBD cases (n=58 donors). Connecting lines indicate paired samples. P values were calculated using non-parametric Kruskal-Wallis (A) or Wilcoxon matched-pairs signed rank tests (B).

### Analysis of *TNF* methylation association with covariates reveals link to anti-TNF therapy response

We next investigated if systemic inflammatory markers were associated with mucosal *TNF* methylation levels. The clinical C-reactive protein (CRP) and erythrocyte sedimentation rate (ESR) values closest to the index colonoscopy when tissue samples were collected demonstrated that CRP and ESR correlated (test of Spearman correlation; CRP vs. *TNF*, p=0.93; ESR vs. *TNF*, p=0.68; ESR vs. CRP, p<0.0001). Among all observations (cases and controls), neither CRP nor ESR significantly correlated with *TNF* methylation, and within cases alone, CRP and ESR also were not correlated with mucosal *TNF* methylation. Some IBD risk exposures, such as tobacco smoke, can have modifying effects on DNA methylation of certain loci^15, 25^. However, neither tobacco nor alcohol use correlated with mucosal *TNF* methylation levels.

We next explored whether mucosal *TNF* methylation levels differed among anti-TNF responders and non-responders. In the initial cohort, the number of non-responders was small with only 5 cases. Therefore, we analyzed our second cohort, which was comprised of a comparable number of responders (15 cases) and non-responders (17 cases, 14 of which were primary non-responders). Strikingly, uninflamed mucosa of non-responders had reduced levels of *TNF* methylation compared to responders (Δ=-9.31 (–12.9%); parametric, unpaired t test, p=0.0359) (Fig. 4A). Non-responders had similar levels of *TNF* methylation in uninflamed and inflamed mucosa (Fig. 4A, B). These results contrast with those of anti-TNF responders, in which *TNF* methylation values strongly correlated with inflammation status (Fig. 2, 3).

**Figure 4.**
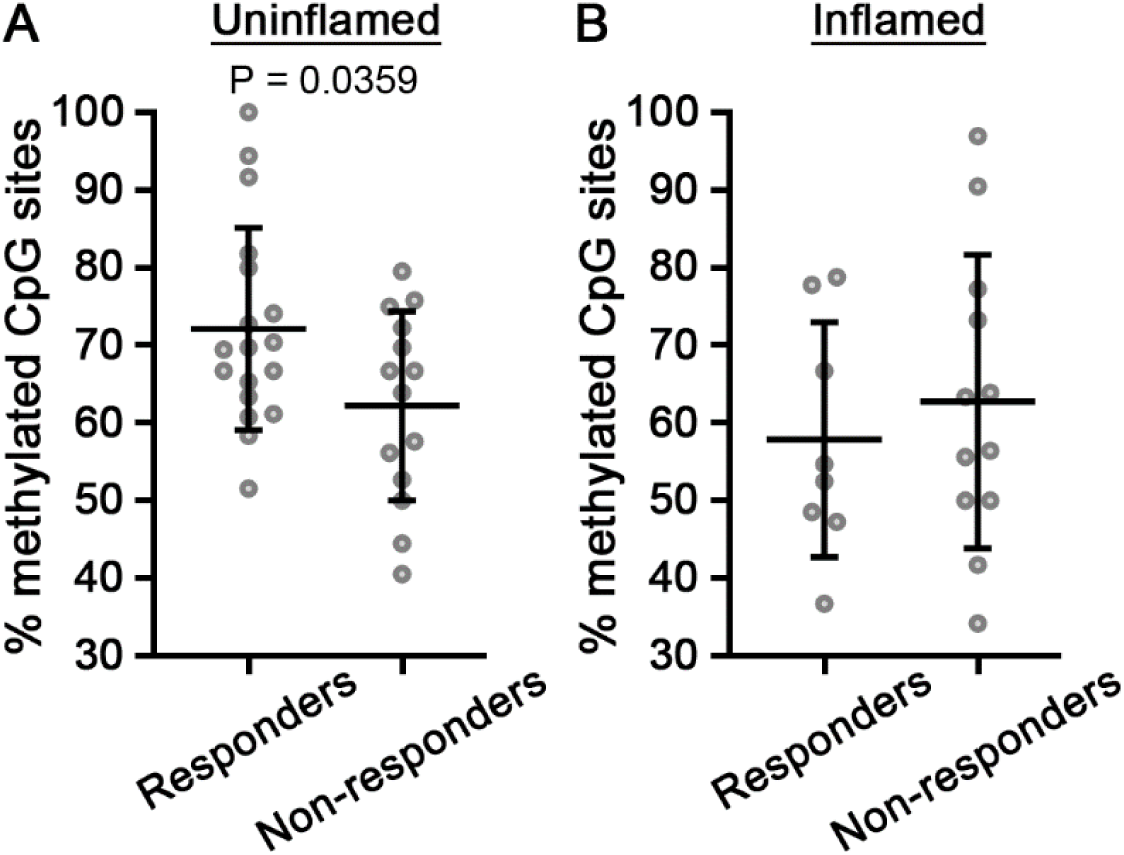
Anti-TNF non-responders have lower levels of *TNF* methylation than responders. (**A-B**) ***TNF*** Methylation analysis of anti-TNF responders and non-responders in uninflamed (A) or inflamed (B) mucosa. Data points are average methylation values from individual cases. Mean ± SD are plotted. P values were calculated using parametric unpaired t tests.

### Contribution of the epithelium to TNF methylation and mucosal inflammation

To investigate the possibility that the methylation differences we observed in inflamed mucosa could derive from changes in cell composition, we tested whether methylation changes within intestinal epithelial cells (IECs) can account for any of the changes we observe in mucosal biopsies. To this end, we isolated IECs using fluorescence activated cell sorting (FACS) from sigmoid or ascending colon bowel resections of non-IBD (NIBD) controls or inflamed CD cases. While methylation levels of both controls and cases were higher in IECs relative to those of mucosal biopsies, IECs isolated from inflamed tissue were hypomethylated compared to control IECs (non-parametric, Wilcoxon rank sum test, p=0.0368) (Fig. 5A), and IECs isolated from inflamed tissue showed greater changes in *TNF* methylation than inflamed mucosa compared to their respective controls (IECs, Δ=-13.87 (–15.2%); mucosa, Δ=-9.09 (–12.3%)). These findings indicate that at least some of the inflammatory changes in mucosal *TNF* methylation are due to changes in IECs themselves rather than to recruitment of inflammatory cell types with diverse methylation levels.

**Figure 5.**
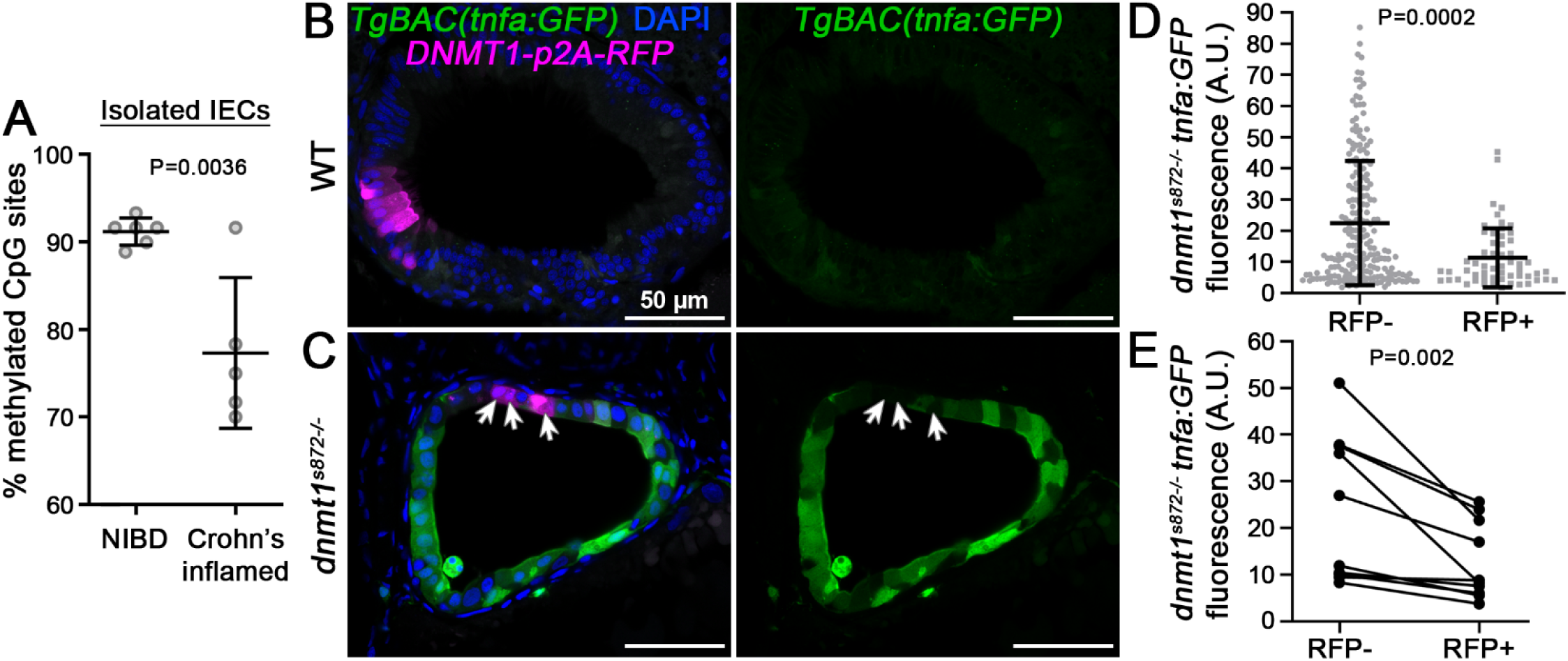
*TNF* hypomethylation of IECs is associated with inflammation and increased IEC *tnf* expression. (**A**) *TNF* Methylation analysis of FACS isolated IECs obtained from bowel resections of non-IBD (NIBD) controls and inflamed CD cases. Data points are average methylation values from individual cases and controls. Mean ± SD are plotted. **(B-C)** 1-cell stage *dnmt1^s872^* zebrafish mutants and WT siblings expressing *TgBAC(tnfa:GFP)^pd1028^* as an inflammation reporter were injected with transgenic constructs to mosaically express human *DNMT1-p2A-RFP* specifically in IECs. At 5 days post fertilization (dpf), transverse sections of the intestine were collected and imaged by confocal microscopy. Arrows point to sparsely labeled RFP+ cells expressing *DNMT1* in *dnmt1^s872^* mutants. Scale bars are 50 µm. **(D)** Quantification of *tnfa:GFP* expression in *dnmt1^s872^*mutants. N=217 (RFP–) and 57 (RFP+) IECs from 10 larvae. Mean ± SD are plotted. **(E)** Quantification of *tnfa:GFP* expression in *dnmt1^s872^*mutants from paired RFP– and RFP+ IECs from individual tissue sections. N=10 larvae. P values were calculated using non-parametric Wilcoxon two sample tests (A, D) or a Wilcoxon matched-pairs signed rank tests (E).

To further explore the role of the epithelium in this process, we turned to a genetic model of spontaneous intestinal inflammation. Zebrafish mutants for the DNA methyltransferase gene, *dnmt1*, develop hallmarks of inflammation in the gut (i.e., cytokine induction, immune cell infiltration, loss of barrier function) in early larval stages^21^. Furthermore, upregulation of *tnf* within mutant IECs was associated with *tnf* promoter hypomethylation and was found to immediately precede subsequent inflammatory phenotypes in *dnmt1* mutants^21^.

To test whether Dnmt1 activity in IECs is sufficient to suppress inflammation, we performed rescue experiments by mosaically expressing human *DNMT1* in small clones of IECs in *dnmt1* mutants, using p2A-mCherry as a reporter for transgenic *DNMT1* expression. In this scenario, mutants are devoid of maintenance DNA methylation except for the sparsely labeled IECs that express human *DNMT1*. To monitor *tnf* expression, we used a transgenic bacterial artificial chromosome (BAC) reporter line we previously generated^21^ and quantified *tnfa:GFP* levels in *DNMT1-p2a-mCherry*+ IECs, using neighboring mutant IECs as internal controls (Fig. 5B, C). We found that mutant IECs expressing *DNMT1* showed a 49.7% reduction in *tnfa:GFP* expression relative to *DNMT1*-negative controls (non-parametric, Wilcoxon rank sum test, p=0.0002) (Fig. 5B, C). Paired *DNMT1*-positive and –negative IECs from individual tissue sections showed similar changes (non-parametric, Wilcoxon matched-pairs signed rank test, p=0.002) (Fig. 5D). *DNMT1*-negative IECs immediately adjacent to *DNMT1*-positive clones had numerically lower levels of *tnfa:GFP* than more distant IECs, although the differences did not reach statistical significance (parametric, paired t test, p=0.0772). These results suggest that DNA methylation within IECs influences local *tnf* expression levels.

### Identification of candidate mutations in *DNMT1* from IBD patients

To investigate whether variants in methyltransferase genes could account for any changes in *TNF* methylation we observed in our cohort, we conducted bidirectional Sanger sequencing of the coding exons and intron-exon junctions in *DNMT1, DNMT3A,* and *UHRF1* in genomic DNA extracted from mucosal biopsies of 27 controls and 70 cases. We identified 2 rare missense variants of interest in *DNMT1* (NM_001130823.3: c.229G>A; p.G77S, and c.4428T>G; p. H1476Q) that were present in separate cases exhibiting *TNF* hypomethylation but were not present in controls and were rare in publicly available reference datasets (genome aggregation database; gnomAD v2.1.1). The G77S variant is evolutionarily constrained and predicted to be pathogenic by two different in silico prediction algorithms (PolyPhen-2, probably damaging; Mutation Taster, disease causing). On the other hand, the H1476Q variant is not predicted to be pathogenic, although it affects the target recognition domain within the methyltransferase domain of DNMT1. We tested if the function of these variants is impaired using rescue experiments in zebrafish. We injected copy RNA (cRNA) encoding WT, G77S, or H1476Q *DNMT1* into crosses of *dnmt1* heterozygotes and then quantified *tnfa:GFP* expression levels in mutant and WT sibling (controls) IECs. Expression of WT *DNMT1* in mutants strongly suppressed *tnfa:GFP* to levels similar to that of WT control siblings (Fig. 6A-C, F). The G77S variant also showed relatively strong activity and partially rescued *tnfa:GFP* expression (Fig. 6B, D, F). By contrast, the H1476Q variant showed significantly lower activity and failed to rescue *tnfa:GFP* levels (Fig. 6B, E, F).

**Figure 6.**
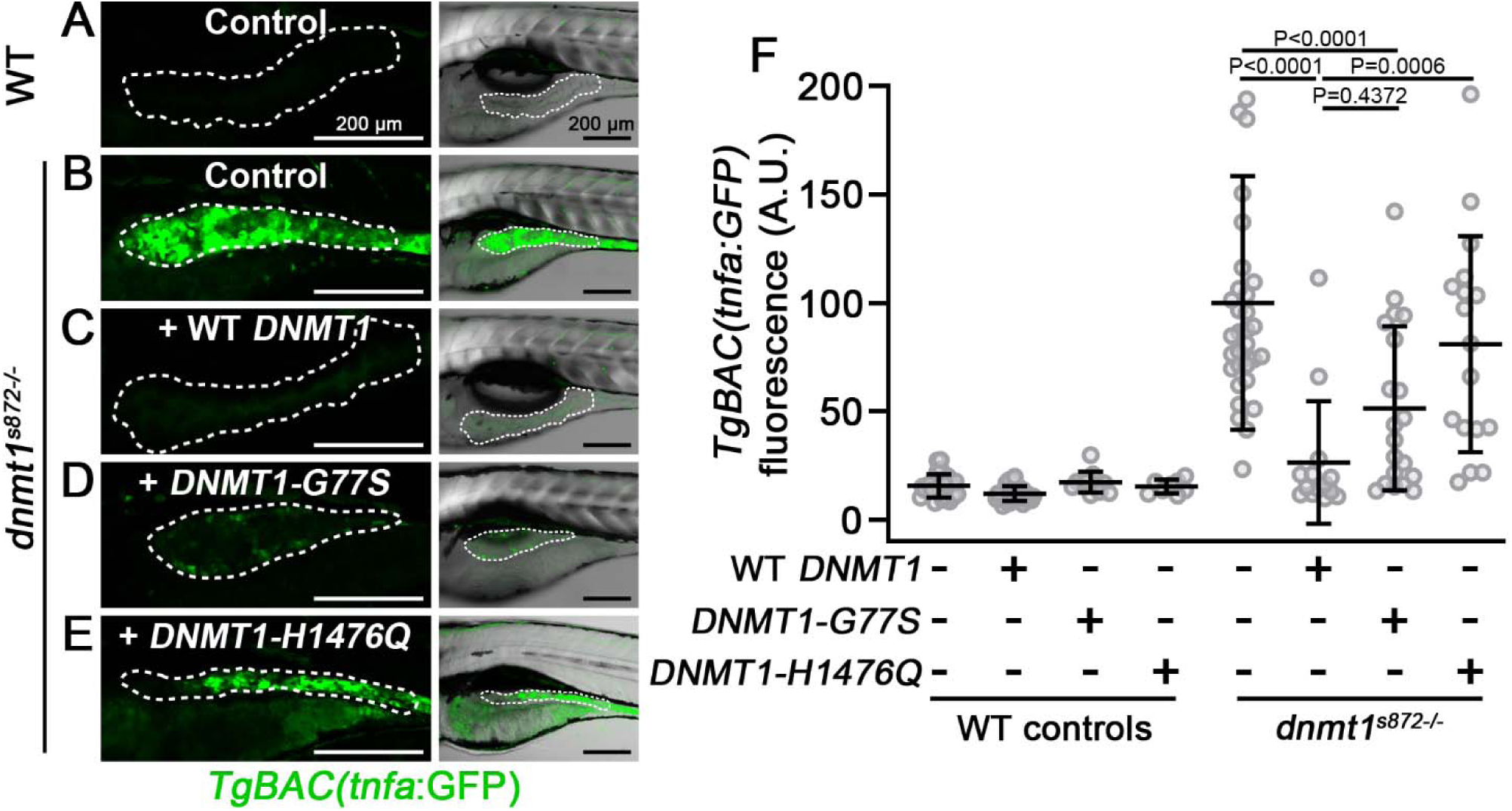
Functional analysis of rare *DNMT1* variants in a zebrafish *dnmt1* mutant model. (**A-E**) Live confocal imaging of *TgBAC(tnfa:GFP)* in 5 dpf *dnmt1^s872^*mutant larvae and WT siblings. 1-cell stage embryos were injected with cRNA encoding WT or variant DNMT1, raised to 5 dpf, imaged, and then genotyped. Dotted line demarcates the anterior intestinal epithelium. Right panels show GFP overlayed with brightfield images. Scale bars are 200 µm. **(F)** Quantification of *tnfa*:GFP intensity in the anterior intestinal epithelium from experiments in panels A-E. Mean ± SD are plotted. P values were calculated using two-way ANOVA. Data points are mean pixel intensity values from individual larvae.

## Discussion

IBD is a complex disorder in which environmental triggers lead to an uncontrolled immune response in the intestine. Although hundreds of genes can confer risk to IBD, inflammation is largely propagated by increased expression of a handful of cytokines, most notably TNF^4, 5^. Given the central role of TNF and the strong influence of environmental factors on IBD, we investigated how epigenetic marks of the *TNF* locus are affected in the mucosa of IBD patients. We found that *TNF* promoter methylation is decreased within inflamed mucosa and IECs isolated from inflamed tissue. Mucosal hypomethylation does not correlate with systemic inflammatory markers and, even among patients with severe active IBD, uninflamed mucosal segments show largely normal levels of *TNF* methylation. We also found that *TNF* hypomethylation correlates with the severity of local inflammation. We discovered that anti-TNF non-responders show distinct patterns of *TNF* methylation, with inflamed and uninflamed segments alike exhibiting hypomethylation. Altogether, our study identifies an association of *TNF* promoter methylation with inflammation in IBD and reveals a novel biomarker that may help to distinguish anti-TNF non-responders from responders.

Our studies using paired inflamed and uninflamed biopsies from individual patients suggest that *TNF* hypomethylation is a consequence of local mucosal inflammation. This conclusion is supported by a recent study showing that inflamed mucosa of IBD patients has reduced expression of DNA methyltransferase genes^23^, and mouse intestinal organoids treated with TNF strongly downregulate *Dnmt3a*^23^. On the other hand, our rescue experiments using zebrafish *dnmt1* mutants indicate that DNA methylation may have cell-autonomous protective effects in an inflammatory microenvironment. Together, these studies suggest that impaired DNA methylation may trigger a feedback loop that exacerbates cytokine expression and inflammation. However, our DNMT1, DNMT3A, and UHRF1 sequencing studies indicate that rare variants in methyltransferase genes are not a predominant contributor impaired DNA methylation in IBD. It would be informative to explore whether DNA methylation in IBD patients is more sensitive to environmental insults, such as infection or diet-associated inflammation.

A key problem in IBD management is that personalized therapy approaches are not currently part of clinical practice. Therapeutic agents are selected empirically rather than in response to a patient’s specific biology. Although anti-TNF therapy remains the most effective treatment for achieving clinical remission and mucosal healing in IBD patients^6^, approximately one third of IBD patients show poor clinical response to anti-TNFs. Because of the incomplete response rate, adverse side effects, and high costs, there is urgent need for markers that can predict a patient’s response to anti-TNF therapy before beginning treatment^26^. We found that uninflamed mucosa of anti-TNF non-responders show reduced levels of *TNF* methylation relative to that of responders. These findings suggest that *TNF* methylation is a quantitative parameter that can be measured pre-therapeutically and could therefore inform the potential for anti-TNF response. Larger, prospective studies examining *TNF* methylation in inflamed and uninflamed tissues of IBD patients before anti-TNF treatment could be helpful to establish *TNF* methylation status as a biomarker to predict anti-TNF response.

Our finding that *TNF* methylation status is correlated with anti-TNF response raises the possibility that *TNF* hypomethylation could influence the response to anti-TNF therapy. Poor response to anti-TNF therapy can, in part, stem from elevated mucosal TNF levels that exceed the concentration of locally available anti-TNF agents^27^. It is conceivable that widespread *TNF* hypomethylation in both inflamed and uninflamed tissue of non-responders may sensitize these patients to heightened TNF expression that overwhelms anti-TNF antibody levels. Future studies investigating how mucosal *TNF* promoter methylation relates to local TNF levels will help to describe its potential role in anti-TNF response. On the other hand, anti-TNF non-response can also stem from immunogenicity or an underlying biological mechanism independent of TNF. As a result, several biologics and small molecule inhibitors that target other pathways have been evaluated for treatment of anti-TNF non-responders^28^. While it is unclear how *TNF* methylation relates to the efficacy of these alternative IBD therapeutics, such studies may help to devise personalized approaches for IBD management.

Treatment decisions for IBD are made through assessment of disease activity, which is monitored using scoring indices encompassing clinical symptoms, systemic biomarkers, and endoscopic observations^29^. Clinical and endoscopic indices vary widely, and many have been validated for CD and UC^29^. However, histologic assessment may provide a more accurate indicator of disease activity, mucosal healing, and potential relapse^30, 31^. Nonetheless, standardized indices for measuring histologic activity of IBD are lacking, particularly for CD^30, 32^. Incorporating molecular biomarkers, such as *TNF* promoter methylation, may aid in the development of an objective scoring index of histologic-level disease activity in CD and UC.

While the differences in *TNF* methylation we observed were robust, prior studies, including genome wide analyses^10–18^, did not identify *TNF* as differentially methylated in IBD. A likely reason that *TNF* was not identified in genome-wide methylation studies of IBD is that the chip arrays used for such analyses, including the latest EPIC BeadChip DNA methylation array^20^, do not include the CpG sites we selected for our study. While genome-wide methylation studies have been instrumental in identifying potential biomarkers for distinguishing IBD cases, our study highlights the utility of targeted methylation studies of IBD candidate genes that may not be captured by commercial arrays.

One limitation of this study is the modest samples sizes in some sub-groups. Although our study included 133 cases and 67 controls, stratifying cases to compare methylation changes with disease activity or location resulted in smaller sample sizes. Similarly, our sample sizes for analyzing methylation changes between anti-TNF responders and non-responders were also small.

Another limitation of this study is that we did not fully define the cellular source(s) of *TNF* hypomethylation in inflamed mucosa from IBD patients and in mucosa from anti-TNF non-responders. IECs represent a major cell type in mucosal biopsies, and our analysis of IECs isolated from inflamed IBD mucosa and healthy donors demonstrated significant methylation differences that were proportional to those of complete mucosal biopsies. This finding supports the hypothesis that IECs are an important source of *TNF* hypomethylation in inflamed IBD mucosa. However, our studies do not exclude the possibility that some of the methylation differences we observed derive from changes in cellular composition. DNA methylation signatures vary among different cell lineages^33^, and biopsies from acutely inflamed mucosa have increased neutrophils and plasma cells. Thus, it is possible that some of the *TNF* methylation differences we observed could be related to increased proportions of differentially methylated cell types and not entirely due to hypomethylation within IECs.

In conclusion, our study reveals an association between *TNF* promoter methylation and mucosal inflammation in CD and UC patients. IECs represent a major source of these *TNF* methylation differences: IECs isolated from CD patients show significantly less *TNF* methylation than IECs from healthy controls, and clinically relevant variants in *DNMT1* influence *tnf* expression in IECs in a vertebrate model system. This study lays the foundation for developing a quantitative biomarker based on *TNF* methylation to help predict anti-TNF therapy responses.

## Data Availability

All data produced in the present study are available upon reasonable request to the authors.

## Abbreviations

BAC: Bacterial Artificial Chromosome
CD: Crohn’s Disease
CpG: Cytosine-phosphate-Guanine
cRNA: copy RNA
CRP: C-Reactive Protein
DNMT1/3A/3B: DNA Methyltransferase 1/3A/3B
ESR: Erythrocyte Sedimentation Rate
FACS: Fluorescence Activated Cell Sorting
GFP: Green Fluorescent Protein
IBD: Inflammatory Bowel Disease
IECs: Intestinal Epithelial Cells
NIBD: non-IBD
TNF: Tumor Necrosis Factor
UC: Ulcerative Colitis
UHRF1: Ubiquitin like with PHD and Ring Finger Domains 1

## Acknowledgements

This study was supported by work from the University of North Carolina Translational Pathology Lab, High Throughput Sequencing Facility, and Tissue Genomic Lab which are supported in part by an NCI Center Core Support Grant (5P30CA016080-42). Technical assistance was provided by Jacob Smoot; animal care and aquaculture services were provided by the Duke Z-Core Facility.

## Methods

All authors had access to the study data and had reviewed and approved the final manuscript.

## Patient selection

Patients undergoing colonoscopy and giving informed consent for research were enrolled in the Duke Gastrointestinal (GI) Tissue Repository under an existing IRB protocol (Pro00001662). Patients presenting for screening or diagnostic colonoscopy without any known history of IBD or colon cancer and giving informed consent for research were selected as controls. Patients with IBD were enrolled at the time of either surveillance or diagnostic colonoscopy. Both UC and CD patients were included. IBD patients represented the spectrum of disease activity.

## Sample collection

In addition to clinical biopsies, up to six research biopsies were collected at the time of colonoscopy by research staff present in the endoscopy room. In patients with IBD undergoing clinical biopsies, the research biopsies were paired with clinical biopsies. For each area where research biopsies were obtained, both endoscopic and microscopic disease activity at that specific location were recorded as part of the research record. In screening colonoscopy patients, clinical biopsies were not typically obtained from normal colonic mucosa and research biopsies were obtained from different parts of the colon to allow comparison with IBD samples from different regions of the colon. In addition to colonic biopsies, in some cases and controls, research samples were also collected from the terminal ileum.

A second cohort of patient samples was identified from the Pathology Archives of the University of Utah (IRB 00091019).

## Sample processing

For the first cohort, research samples were collected between 2014 and 2020. Research samples obtained during the index colonoscopy were placed in optimal cutting temperature compound and snap frozen over either liquid nitrogen or dry ice and stored at –80 degrees C. Frozen samples were then processed in Duke’s BioRepository & Precision Pathology Core to obtain DNA for the methylation assays.

For the second cohort, scrolls (mucosal biopsies) or unstained slides (resections) were obtained from formalin-fixed paraffin-embedded tissue blocks. Mucosal areas were microdissected (resections). DNA was prepared by the University of Utah Biorepository and Molecular Pathology Shared Resource.

De-identified DNA samples were processed for bisulfite conversion using the EZ DNA Methylation-Direct kit (Zymo Research), and a *TNF* promoter fragment was amplified using nested PCR with the primers TNF_outer_f 5’-CTAACTAAATATACCAACAACTA-3’ and TNF_outer_r 5’-AGAAATGGAGGTAATAGGTTTT-3’ followed by TNF_inner_f 5’-CCAACAACTACCTTTATATATC-3’ and TNF_inner_r 5’-AGGTTTTGAGGGGTATGGG-3’. PCR amplicons were purified and cloned using the pGEM-T Easy Vector System (Promega). Sanger sequencing of bacterial colonies was performed by Genewiz (Azenta Life Sciences), and sequencing data were analyzed using QUMA^34^.

## Statistical methods

General patient characteristics were described and compared between cases and controls. Age was compared using the Kruskal-Wallis test and sex and race were compared using the Fisher Exact Test. Descriptive statistics were compiled for methylation markers overall. These assessments included multiple observations per subject. The distribution of methylation values did not meet the assumption of normality. Thus, the non-parametric, Wilcoxon Two Sample test, was used to compare methylation values between cases and controls, between inflamed and uninflamed samples, and between anti-TNF responders and non-responders assuming independence among observations. TNF methylation levels by inflammation status (paired) within subject were compared using the Wilcoxon Signed Rank test. Spearman correlation estimates were computed to assess relationships between ESR, CRP and methylation status (results not shown). Analyses were performed in SAS 9.4 and GraphPad Prism 10.0.2.

## Clinical data collection

Clinical data related to IBD were collected using a detailed chart abstraction process from the electronic medical record. Both cases and controls had basic demographic data collected including age at time of sample collection, sex, and race/ethnicity. IBD patients also had data collected including length of time since diagnosis with IBD, type of IBD (CD, UC, indeterminate), type of disease (ileal, colonic, upper GI tract, fistulizing, perianal, proctitis only). Disease activity at the time of sample collection was determined by presence of symptoms, endoscopic disease activity determined by review of the endoscopy report, and histologic evidence of disease determined by pathology report generated by a GI clinical pathologist. Family history of IBD and surgical history were noted. Treatment history was classified based on any prior anti-TNF therapy and, if treated, response to that therapy. Other medication use was recorded. Laboratory data closest to the time of index colonoscopy were recorded including erythrocyte sedimentation rate and c-reactive protein. Exposures such as tobacco and alcohol use were recorded. Clinical recurrence data were also abstracted from the medical record.

## Isolation of intestinal epithelial cells (IECs)

Colonic epithelial cells were isolated as previously described^35, 36^. In brief, dissected colonic mucosa was cut into small pieces and incubated in magnesium-free Hank’s balanced salt solution (HBSS) containing 2 mmol/L EDTA and 2.5% heat-inactivated fetal bovine serum for 30 minutes with shaking at 37°C. Collected supernatants were centrifuged, resuspended in HBSS containing 1 mg/mL collagenase type 4 (17104019; Thermo Fisher Scientific), and incubated for 10 minutes at 37°C to further remove the mucus. The fraction was pelleted, resuspended in HBSS, passed through a 40-µm filter, and overlayered on 50% Percoll. Cells were centrifuged at 2000 rpm for 20 minutes at room temperature and viable colonic IECs were recovered from the interface layer.

## Mutational analysis

Coding exons and intron-exon junctions of *DNMT1*, *DNMT3A*, and *UHRF1* (79 exons total) were PCR-amplified and sequenced bidirectionally using BigDye Terminator 3.1 chemistry on an ABI 3730xl automated capillary sequencer (Applied Biosystems). Sequencher (Gene Codes) was used for sequence alignment to reference. Primer sequences are available upon request. Variant frequency information was obtained from gnomAD (gnomAD.broadinstitute.org); in silico prediction information was queried on PolyPhen-2 (genetics.bwh.harvard.edu/pph2/) and Mutation Taster (mutationtaster.org).

## Zebrafish experiments

Zebrafish (*Danio rerio*) were used in accordance with Duke University Institutional Animal Care and Use Committee (IACUC) guidelines under the approval from protocol number 170105–02. Zebrafish stocks were maintained and bred as previously described^37^. Genotypes were determined by PCR and DNA sequencing or phenotypic analysis. Male and female breeders from 3–18 months of age were used to generate fish for all experiments. Zebrafish larvae (5-6 days post fertilization) from the Ekkwill (EK) background were used in this study. Strains used in this study were: *dnmt1*^*s872* 38^ and *TgBAC(tnfa:GFP)*^*pd1028* 21^. Larvae were anesthetized with 0.4 mg/mL MS-222 (Sigma, A5040) dissolved in embryo media for handling when necessary.

For mosaic transgene expression, the coding sequence of *DNMT1* (NM_001130823) was subcloned to pDONR221 using gateway cloning (Thermo Fisher Scientific). pDEST-Tol2-QUAS:DNMT1-p2A-mCherry and pDEST-Tol2-cldn15la:QF2 were constructed as previously described^39^ and co-injected with transposase copy RNA (cRNA) into one-cell stage embryos generated from crosses of *dnmt1^s872^; TgBAC(tnfa:GFP)^pd1028^* double heterozygotes. Transverse sections of the mid-intestine of mutant and WT sibling larvae were then collected using a Leica VT1000S vibratome as previously described^40^, and confocal imaging was performed with an Olympus Fluoview FV3000 with a 60x/1.4 N.A. oil objective. Confocal data were analyzed using ImageJ/FIJI (National Institutes of Health) and Graphpad Prism 10.0.2.

For rescue experiments, the coding sequence of *DNMT1* (NM_001130823) was subcloned to pCSDest^41^ using gateway cloning (Thermo Fisher Scientific). Variant substitutions were introduced using Q5 site directed mutagenesis (New England Biolabs). *DNMT1* WT or variant cRNA was injected into one-cell stage embryos generated from crosses of *dnmt1^s872^; TgBAC(tnfa:GFP)^pd1028^* double heterozygotes. Live confocal imaging was performed with an Olympus Fluoview FV3000 with a 10x/0.4 N.A. objective. Confocal data were analyzed using ImageJ/FIJI (National Institutes of Health) and then larvae were genotyped before statistical analysis (Graphpad Prism).

## References

1. Jostins L, Ripke S, Weersma RK, et al. Host-microbe interactions have shaped the genetic architecture of inflammatory bowel disease. Nature 2012;491:119–24.

2. Liu JZ, van Sommeren S, Huang H, et al. Association analyses identify 38 susceptibility loci for inflammatory bowel disease and highlight shared genetic risk across populations. Nat Genet 2015;47:979–986.

3. de Lange KM, Moutsianas L, Lee JC, et al. Genome-wide association study implicates immune activation of multiple integrin genes in inflammatory bowel disease. Nat Genet 2017;49:256–261.

4. Neurath MF. Cytokines in inflammatory bowel disease. Nat Rev Immunol 2014;14:329–42.

5. D’Haens GR, van Deventer S. 25 years of anti-TNF treatment for inflammatory bowel disease: lessons from the past and a look to the future. Gut 2021;70:1396–1405.

6. Narula N, Wong ECL, Dulai PS, et al. Comparative Effectiveness of Biologics for Endoscopic Healing of the Ileum and Colon in Crohn’s Disease. Am J Gastroenterol 2022;117:1106–1117.

7. Olsen T, Goll R, Cui G, et al. Tissue levels of tumor necrosis factor-alpha correlates with grade of inflammation in untreated ulcerative colitis. Scand J Gastroenterol 2007;42:1312–20.

8. Stylianou E. Epigenetics of chronic inflammatory diseases. J Inflamm Res 2019;12:1–14.

9. Feil R, Fraga MF. Epigenetics and the environment: emerging patterns and implications. Nat Rev Genet 2012;13:97–109.

10. Cooke J, Zhang H, Greger L, et al. Mucosal genome-wide methylation changes in inflammatory bowel disease. Inflamm Bowel Dis 2012;18:2128–37.

11. Nimmo ER, Prendergast JG, Aldhous MC, et al. Genome-wide methylation profiling in Crohn’s disease identifies altered epigenetic regulation of key host defense mechanisms including the Th17 pathway. Inflamm Bowel Dis 2012;18:889–99.

12. Harris RA, Nagy-Szakal D, Pedersen N, et al. Genome-wide peripheral blood leukocyte DNA methylation microarrays identified a single association with inflammatory bowel diseases. Inflamm Bowel Dis 2012;18:2334–41.

13. Adams AT, Kennedy NA, Hansen R, et al. Two-stage genome-wide methylation profiling in childhood-onset Crohn’s Disease implicates epigenetic alterations at the VMP1/MIR21 and HLA loci. Inflamm Bowel Dis 2014;20:1784–93.

14. Harris RA, Nagy-Szakal D, Mir SA, et al. DNA methylation-associated colonic mucosal immune and defense responses in treatment-naive pediatric ulcerative colitis. Epigenetics 2014;9:1131–7.

15. Ventham NT, Kennedy NA, Adams AT, et al. Integrative epigenome-wide analysis demonstrates that DNA methylation may mediate genetic risk in inflammatory bowel disease. Nat Commun 2016;7:13507.

16. McDermott E, Ryan EJ, Tosetto M, et al. DNA Methylation Profiling in Inflammatory Bowel Disease Provides New Insights into Disease Pathogenesis. J Crohns Colitis 2016;10:77–86.

17. Kraiczy J, Nayak K, Ross A, et al. Assessing DNA methylation in the developing human intestinal epithelium: potential link to inflammatory bowel disease. Mucosal Immunol 2016;9:647–58.

18. Howell KJ, Kraiczy J, Nayak KM, et al. DNA Methylation and Transcription Patterns in Intestinal Epithelial Cells From Pediatric Patients With Inflammatory Bowel Diseases Differentiate Disease Subtypes and Associate With Outcome. Gastroenterology 2018;154:585–598.

19. Hornschuh M, Wirthgen E, Wolfien M, et al. The role of epigenetic modifications for the pathogenesis of Crohn’s disease. Clin Epigenetics 2021;13:108.

20. Joustra V, Li Yim AYF, Hageman I, et al. Long-term Temporal Stability of Peripheral Blood DNA Methylation Profiles in Patients With Inflammatory Bowel Disease. Cell Mol Gastroenterol Hepatol 2023;15:869–885.

21. Marjoram L, Alvers A, Deerhake ME, et al. Epigenetic control of intestinal barrier function and inflammation in zebrafish. Proc Natl Acad Sci U S A 2015;112:2770–5.

22. Elliott EN, Sheaffer KL, Kaestner KH. The ‘de novo’ DNA methyltransferase Dnmt3b compensates the Dnmt1-deficient intestinal epithelium. Elife 2016;5.

23. Fazio A, Bordoni D, Kuiper JWP, et al. DNA methyltransferase 3A controls intestinal epithelial barrier function and regeneration in the colon. Nat Commun 2022;13:6266.

24. Gowers IR, Walters K, Kiss-Toth E, et al. Age-related loss of CpG methylation in the tumour necrosis factor promoter. Cytokine 2011;56:792–7.

25. Tsaprouni LG, Yang TP, Bell J, et al. Cigarette smoking reduces DNA methylation levels at multiple genomic loci but the effect is partially reversible upon cessation. Epigenetics 2014;9:1382–96.

26. Cui G, Fan Q, Li Z, et al. Evaluation of anti-TNF therapeutic response in patients with inflammatory bowel disease: Current and novel biomarkers. EBioMedicine 2021;66:103329.

27. Yarur AJ, Jain A, Hauenstein SI, et al. Higher Adalimumab Levels Are Associated with Histologic and Endoscopic Remission in Patients with Crohn’s Disease and Ulcerative Colitis. Inflamm Bowel Dis 2016;22:409–15.

28. Singh S, Murad MH, Fumery M, et al. Comparative efficacy and safety of biologic therapies for moderate-to-severe Crohn’s disease: a systematic review and network meta-analysis. Lancet Gastroenterol Hepatol 2021;6:1002–1014.

29. Kishi M, Hirai F, Takatsu N, et al. A review on the current status and definitions of activity indices in inflammatory bowel disease: how to use indices for precise evaluation. J Gastroenterol 2022;57:246–266.

30. Vespa E, D’Amico F, Sollai M, et al. Histological Scores in Patients with Inflammatory Bowel Diseases: The State of the Art. J Clin Med 2022;11.

31. Gupta A, Yu A, Peyrin-Biroulet L, et al. Treat to Target: The Role of Histologic Healing in Inflammatory Bowel Diseases: A Systematic Review and Meta-analysis. Clin Gastroenterol Hepatol 2021;19:1800–1813 e4.

32. Novak G, Parker CE, Pai RK, et al. Histologic scoring indices for evaluation of disease activity in Crohn’s disease. Cochrane Database Syst Rev 2017;7:CD012351.

33. Loyfer N, Magenheim J, Peretz A, et al. A DNA methylation atlas of normal human cell types. Nature 2023;613:355–364.

34. Kumaki Y, Oda M, Okano M. QUMA: quantification tool for methylation analysis. Nucleic Acids Res 2008;36:W170–5.

35. Keith BP, Barrow JB, Toyonaga T, et al. Colonic epithelial miR-31 associates with the development of Crohn’s phenotypes. JCI Insight 2018;3.

36. Toyonaga T, Steinbach EC, Keith BP, et al. Decreased Colonic Activin Receptor-Like Kinase 1 Disrupts Epithelial Barrier Integrity in Patients With Crohn’s Disease. Cell Mol Gastroenterol Hepatol 2020;10:779–796.

37. Westerfield M. THE ZEBRAFISH BOOK, 5th Edition; A guide for the laboratory use of zebrafish (Danio rerio): University of Oregon Press, 2007.

38. Anderson RM, Bosch JA, Goll MG, et al. Loss of Dnmt1 catalytic activity reveals multiple roles for DNA methylation during pancreas development and regeneration. Dev Biol 2009;334:213–23.

39. Kwan KM, Fujimoto E, Grabher C, et al. The Tol2kit: a multisite gateway-based construction kit for Tol2 transposon transgenesis constructs. Dev Dyn 2007;236:3088–99.

40. Levic DS, Ryan S, Marjoram L, et al. Distinct roles for luminal acidification in apical protein sorting and trafficking in zebrafish. J Cell Biol 2020;219.

41. Villefranc JA, Amigo J, Lawson ND. Gateway compatible vectors for analysis of gene function in the zebrafish. Dev Dyn 2007;236:3077–87.

